# Relative Importance of Positive and Potentially Traumatic Childhood Experiences in Explaining Multiple Sclerosis Symptoms

**DOI:** 10.64898/2026.10.29.26366800

**Authors:** Caterina Obenauf, Alejandro L. Vázquez, Gina P. Owens

**Affiliations:** Department of Psychology and Neuroscience, The University of Tennessee, Knoxville, TN; Department of Rehabilitation Medicine, Emory University School of Medicine, Atlanta, GA

**Author notes:** **Corresponding Author:** Please send general questions about the study to Caterina Obenauf., Caterina Obenauf, M.A., 1404 Circle Dr, Knoxville, TN 37916. **Data Availability Statement:** Due to the sensitive nature of the data in this study (i.e., childhood and present participant zip codes), data from the current study will only be shared upon request at the discretion of the corresponding author. **Ethical Standards Statement:** This study protocol was reviewed and approved by the University of Tennessee Human Research Protections Program, approval number UTK IRB-23-07938-XM.

**Keywords:** ACEs, positive childhood experiences, multiple sclerosis, cognition, fatigue, pain, depression

## Abstract

The original adverse childhood experiences study linked childhood experiences to health in adulthood. Since then, other traumatic and adverse childhood experiences (TRACEs) and positive childhood experiences (PCEs) have also been linked to physical and mental health. However, little research has examined these experiences among people with Multiple Sclerosis (MS). This study examined whether cumulative and individual childhood experiences explained variability in symptom burden in adults with MS. Participants were 467 adults with MS (Mage = 41.5, 45% female) who completed measures of pain, subjective cognitive impairment, cognitive fatigue, depression, TRACEs, and PCEs. The elastic net algorithm examined cumulative TRACEs, PCEs, their interaction, and individual childhood experiences. R-squared was used to assess the proportion of variance in each outcome accounted for by childhood experiences, and variable importance identified experiences that most improved the model’s explanatory power for each outcome. Individual childhood experiences accounted for 26.0%, 17.1%, 13.9%, and 22.4% of variance in pain, subjective cognitive impairment, cognitive fatigue, and depression, respectively, and overall had stronger performance relative to the cumulative main effects TRACEs and PCEs model, and their interaction. Having at least one safe caregiver was consistently important across MS symptoms, and sudden loss of a loved one had the strongest explanatory association with subjective cognitive impairment and cognitive fatigue specifically. Specific childhood experiences accounted for meaningful variance in the MS symptom burden. Individual childhood experiences provided more explanatory information than cumulative scores. Findings support examining specific TRACEs and PCEs when investigating symptom burden in MS.

---

Multiple Sclerosis (MS) is a chronic autoimmune disease that affects the central nervous system, causing symptoms across several health domains that contribute to disability. The combined burden of varied symptoms, such as cognitive impairment, fatigue, pain, and depression, negatively impacts quality of life [1]. MS symptoms and disability outcomes are shaped by both biological disease processes and early-life experiences [2].

Adverse childhood experiences (ACEs) have been extensively linked to poor health conditions in adulthood [3,4]; however, the relationship between ACEs and MS remains unclear and understudied [1,2]. Investigation into this relationship is important because inflammation is implicated both as a central driver of neuronal damage in MS and as a mechanism linking early adversity to poor health in adulthood [5,6]. Across studies, childhood adversity has been associated with increased MS risk, earlier disease onset, higher relapse rates, and greater symptom severity, with emotional abuse and neglect emerging as particularly consistent correlates, in some cases more than tripling the odds of an MS diagnosis [2,7]. However, effects vary by type of ACE, with most studies emphasizing cumulative ACE exposure rather than specific experiences [2]. Thus, cumulative scores may obscure meaningful differences in how particular adversities relate to MS.

Two limitations of the original ACEs framework [3] are especially relevant to MS. First, the original list of ACEs omitted community-level adversities, such as peer victimization and neighborhood violence, that have been linked to poor health in adulthood [8]. Second, the original framework did not incorporate several Criterion A traumatic events, despite evidence that such events similarly affect health as the original ACEs [9]. To address these gaps, Weems and colleagues [10] proposed the Traumatic and Adverse Childhood Experiences (TRACEs) framework, which integrates expanded ACEs [8] with Criterion A events. Because TRACEs are grounded in neuroscientific research linking childhood adversity to brain structure and function, it is particularly well-suited as a theoretical framework in which to study the effects of ACEs on MS; however, to date, the TRACEs framework has not been applied to MS.

Although biological and socioeconomic protective factors in MS have received considerable attention (e.g., vitamin D, educational attainment) [11,12], positive childhood experiences (PCEs), such as having a supportive friend or trusted adult [13], remain largely unexamined. PCEs and ACEs frequently co-occur, and PCEs can buffer the adverse effects of childhood adversity [14], but MS research has remained largely deficit-focused. Considering PCEs and TRACEs simultaneously is consistent with the TRACEs framework’s nuanced integration of risk and resilience [10], and may better capture heterogeneity of symptoms among people with MS.

The current study had two aims. First, we characterized the relative importance of cumulative TRACEs and PCEs, and their interaction, in explaining pain, subjective cognitive impairment, cognitive fatigue, and depressive symptoms in MS. Second, we characterized which individual TRACEs and PCEs accounted for the greatest proportion of variance in those same MS symptoms. Because little research has examined specific TRACEs and PCEs in MS, we did not advance a priori hypotheses regarding which cumulative and individual experiences would show the strongest associations with each outcome.

## Method

### Participants and Procedure

Participants were 467 individuals from a broader study on MS quality of life who reported an MS diagnosis (Mage = 41.52, SD = 12.16, range 19-84; 85% white, 85% heterosexual, 43% cisgender women; [Redacted for Blind Review]). Almost half of participants had relapsing-remitting MS (49%), followed by primary progressive (27%), secondary progressive (25%), and clinically isolated syndrome (5%). Educational attainment was high in this sample (11% completed a graduate degree, 31% completed an undergraduate or vocational degree, 22% completed some post-secondary education, 27% completed high school, and 7% did not complete high school).

Additional demographic information is reported (Supplemental Table S1). Participants were recruited from ResearchMatch, NARCOMS Registry for MS, iConquerMS, and the National MS Society newsletter and website between 27/03/2024 and 27/08/2024. Participant confidentiality and data privacy were ensured throughout the study, and participants provided informed consent before participation, in compliance with ethical guidelines regarding human subjects research and the Declaration of Helsinki. Participants completed a screening survey to ensure they met inclusion criteria: 18 years old or older, diagnosis of MS, and United States residence. Participants were compensated for their participation with a retail gift card. Study procedures were reviewed and approved by the local Institutional Review Board.

### Measures

#### Traumatic and Adverse Childhood Experiences

The TRACES questionnaire [10] includes the 10-item ACEs questionnaire [3] and 10 additional items [8,10]. Additional items either overlap with the DSM-5 criteria for a Criterion A traumatic event or have much evidence to suggest their inclusion with ACEs measures (e.g., racial/ethnic discrimination, separation/loss of a loved one) [10]. Participants were asked to respond to whether they had experienced any of the above items before the age of 18 using a dichotomous scale (Yes/No). The sum of affirmative responses provides a measure of participants’ cumulative TRACE score, with higher scores representing greater adversity exposure. A previous study utilizing this questionnaire indicated strong construct and concurrent validity with other measures of ACEs, and predictive validity for psychopathology such as depression symptoms [15].

#### Positive Childhood Experiences

The Benevolent Childhood Experiences (BCE) Scale is a 10-item scale which measures positive childhood experiences that occurred before the age of 18 [13]. The BCE scale was developed as a counterpart to the ACEs questionnaire and is intended to be used with adults who also have histories of ACEs or other traumatic early life experiences [13]. The BCE scale demonstrates high test-retest reliability, cross-cultural validity among various racial/ethnic and linguistic groups, and predictive validity for psychopathology and stress in adulthood [13].

#### Pain

The Pain Effects Scale (PES) [16] is a 6-item scale that measures the degree to which pain has disrupted mood, mobility, sleep, work, leisure activities, and overall quality of life. The PES has previously demonstrated high internal consistency for the scale (Cronbach’s alpha = .86) in a clinical sample of MS patients [16]. In the current study, Cronbach’s alpha was .84.

#### Subjective Cognitive Impairment

The Perceived Deficits Questionnaire (PDQ) [16] is a 20-item subjective self-report measure of subjective cognitive impairment in the context of MS. The PDQ assesses specific cognitive domains affected by MS (attention, retrospective memory, prospective memory, and planning/organization), with greater scores indicating greater perceived cognitive impairment. The PDQ demonstrates good internal consistency reliability with people with MS (Cronbach’s alpha = .93) [16]. In the current study, Cronbach’s alpha was .92.

#### Cognitive Fatigue

The cognitive fatigue subscale of the Modified Fatigue Impact Scale (MFIS) [17] is a 10-item scale that assesses the impact of cognitive fatigue on daily life in an MS context. The MFIS has previously demonstrated high internal consistency for the cognitive subscale (Cronbach’s alpha = .95), in addition to discriminant validity with other causes of fatigue (e.g., chronic fatigue) in a clinical sample of MS patients [16]. In the current study, the cognitive fatigue subscale of the MFIS had a Cronbach’s alpha of .88.

#### Depression

The depression subscale of the Mental Health Inventory [16] is a 5-item scale that measures depression symptoms in the past four weeks. The Mental Health Inventory and its subscales have convergent validity with other measures of these constructs, and the total scale and depression subscale have good internal consistency reliability in a clinical sample of MS patients (Cronbach’s alpha = .93 and .87, respectively) [16]. In the current study, the depression subscale had a Cronbach’s alpha of .77.

### Analytic Plan

Analyses were performed in R Version 2026 [18]. Regarding pattern of missing data, 44.0% of participants were missing at least one item, with 11.3% missing a single item across all variables, 3.4% missing two items, and 1.5% missing three or more items. However, there was low overall missing data (0.4%). Frequencies and percentages of missing data for individual variables of interest are reported (Supplemental Table S2). We performed a chi-square test of independence to identify a potential pattern of missingness. Participant responses across self-report measures were significantly (p < .05) related to propensity for missingness, with participants reporting greater attention deficits and positive and potentially traumatic childhood experiences being associated with missing data; thus, the pattern of missing data was not missing completely at random. Accordingly, missing values were imputed at the item level using the random forest algorithm, as listwise deletion may introduce additional biases [19,20].

We used a machine learning approach to accommodate the relatively large number of correlated predictors. This method uses cross-validation to reduce overfitting and improve generalizability and identifies variables that meaningfully contribute to each dependent variable [21]. Separate elastic-net penalized linear regression models were fitted to identify TRACEs and PCEs in childhood associated with severity of MS symptoms, including pain, subjective cognitive impairment, cognitive fatigue, and depression symptoms. Elastic net regularization produces more parsimonious models while allowing correlated independent variables that offer unique predictive information to be retained [21]. We conducted 1,000 bootstrap resamples to evaluate model performance and estimate the relative importance of each childhood experience in predicting each MS symptom outcome [22]. We also evaluated the relative importance of cumulative TRACEs and PCEs and the interaction between TRACEs and PCEs. Higher variable-importance values indicated that an independent variable made a larger relative contribution to explaining variability in the dependent variable, compared with the other predictors considered. Model performance for each dependent variable was evaluated using *R*^2^, which represents the proportion of variance in the dependent variable explained by the variables within the model [23].

## Results

Almost all participants (93.9%) endorsed at least one TRACE, most commonly experiencing a lack of neighborhood safety (52%), natural disaster (41%), bullying (41%), separation or sudden loss of a family member or close friend (40%), and verbal abuse (37%). All participants endorsed at least one PCE, most commonly feeling comfortable with themselves (39%), liking school (36%), having opportunities for enjoyment (35%), having at least one supportive non-parental adult figure (34%), and having at least one caring teacher (34%). On average, participants were likely to endorse moderate severity of depression symptoms (mean item response = 3.1/6, SD = 1), subjective cognitive impairment (1.8/4, SD = .62), and cognitive fatigue (1.8/4, SD = .68), and mild impairment from pain across activities of daily living (1.8, SD = .80). Means and standard deviations are presented (Table 1).

**Table 1.** Correlations Among Variables of Interest.

| Variable | Mean | SD | 2 | 3 | 4 | 5 | 6 | 7 |
| --- | --- | --- | --- | --- | --- | --- | --- | --- |
| 1. Traumatic and Adverse Childhood Experiences | 5.97 | 3.80 | .904*** | -.529*** | .187*** | .156*** | .232*** | .304*** |
| 2. Adverse Childhood Experiences | 2.86 | 2.32 | — | -.509*** | .095** | .011 | .133*** | .230*** |
| 3. Positive Childhood Experiences | 6.87 | 2.63 |  | — | .017 | .043 | -.001 | -.259*** |
| 4. Cognitive Fatigue | 17.92 | 6.83 |  |  | — | .504*** | .781*** | .398*** |
| 5. Pain | 10.98 | 4.82 |  |  |  | — | .528*** | .396*** |
| 6. Subjective Cognitive Impairment | 35.06 | 12.47 |  |  |  |  | — | .431*** |
| 7. Depression | 12.46 | 4.06 |  |  |  |  |  | — |

*Note.* \* $p < .05$ . \*\* $p < .01$ . \*\*\* $p < .001$ .

### Associations Between Childhood Experiences and MS Symptoms

Experiencing more TRACEs was associated with experiencing fewer positive childhood experiences, greater pain, subjective cognitive impairment, cognitive fatigue, and depression (*p* < .001). Experiencing more PCEs was associated with experiencing less depression *(p* < .001), but PCEs were not significantly associated with pain, subjective cognitive impairment, or cognitive fatigue (*p*s > .05). The correlation matrix is presented (Table 1).

Regarding the associations among TRACEs and MS symptoms, greater cognitive fatigue was mildly associated with many TRACEs, with the strongest associations with verbal abuse and emotional neglect (ps < .001). Greater pain was associated with several TRACEs, with the strongest associations with racial/ethnic discrimination and sudden loss of a loved one (ps < .001). Greater subjective cognitive impairment was associated with several TRACEs, with the strongest associations with verbal abuse, emotional neglect, and physical neglect (ps < .001). Greater depression was associated with several TRACEs, with the strongest associations with emotional and physical neglect (ps < .001).

Regarding PCEs, having at least one safe caregiver was consistently associated with fewer MS symptoms overall (p < .001). Depression was negatively associated with many other PCEs, with the strongest associations with opportunities for enjoyment and feeling comfortable with oneself. Correlations with individual TRACEs and PCEs are visually presented (Supplemental Figure S1).

### Relative Importance of Individual Childhood Experiences and MS Symptoms

Relative importance of childhood experiences in their association with MS symptoms is presented (Figure 1). Childhood experiences accounted for 26.0% of the variance in pain. Having at least one safe caregiver (PCE) had the highest importance in explaining the variance in pain scores, followed by racial/ethnic discrimination (TRACE), sudden loss of a loved one (TRACE), natural disaster exposure (TRACE), mental illness in the home (TRACE), and physical abuse (TRACE).

**Figure 1.**
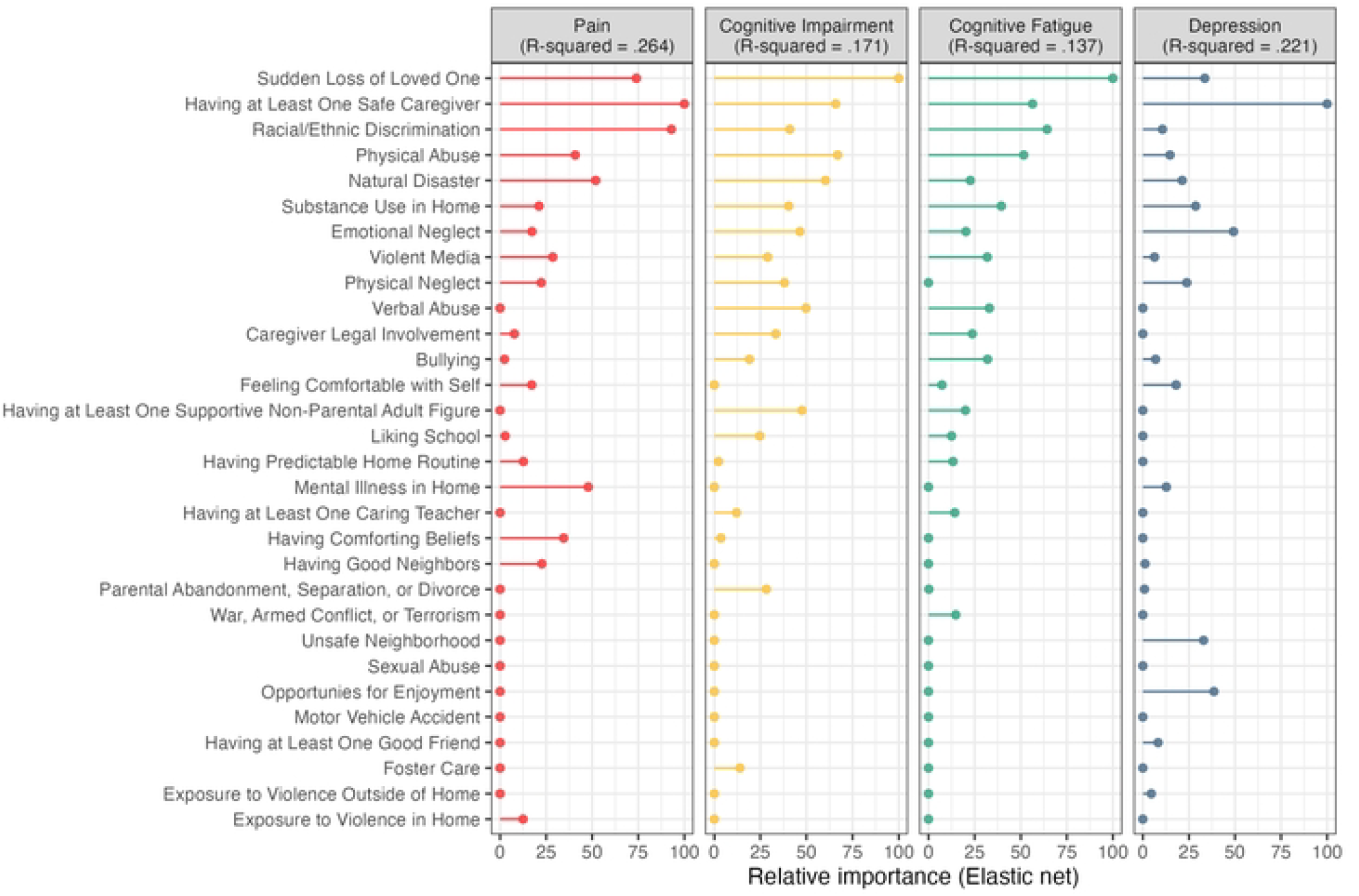
The Relative Importance of Childhood Experiences in Explaining the Variance in Multiple Sclerosis Symptoms

Childhood experiences accounted for 17.1% of the variance in subjective cognitive impairment. Sudden loss of a loved one had the highest importance (TRACE) in explaining the variance in subjective cognitive impairment scores, followed by physical abuse (TRACE), having at least one safe caregiver (PCE), natural disaster exposure (TRACE), verbal abuse (TRACE), and having at least one supportive non-parental adult figure (PCE).

Childhood experiences accounted for 13.9% of the variance in cognitive fatigue. Sudden loss of a loved one had the highest importance (TRACE) in explaining the variance in cognitive fatigue scores, followed by racial/ethnic discrimination (TRACE), having at least one safe caregiver (PCE), physical abuse (TRACE;), and substance use in the home (TRACE).

Childhood experiences accounted for 22.4% of the variance in depression. Having at least one safe caregiver had the highest importance (PCE) in explaining the variance in depression scores, followed by emotional neglect (TRACE), opportunities for enjoyment (TRACE, unsafe neighborhood (TRACE), and sudden loss of a loved one (TRACE).

### Relative Importance of Cumulative Childhood Experiences and MS Symptoms

In contrast, cumulative TRACEs and PCEs accounted for only 3.3% of the variance in pain, 7.3% of the variance in subjective cognitive impairment, 5.5% of the variance in cognitive fatigue, and 10.4% of the variance in depression. Cumulative TRACEs accounted for the variance in pain, subjective cognitive impairment, and cognitive fatigue according to variable importance, and cumulative PCEs had no variable importance in improving model performance for these outcomes. In contrast, cumulative PCEs accounted for the observed variance in depression according to variable importance, whereas cumulative TRACEs had no variable importance in accounting for this outcome. The TRACE x PCE interaction yielded lower variance explained than the main effects (*R*^2^ pain = .037; *R*^2^ subjective cognitive impairment = .069; *R*^2^ cognitive fatigue = .051; *R*^2^ depression = .103) compared to the cumulative model. Overall, models incorporating individual childhood experiences demonstrated greater performance than models using cumulative TRACEs and PCEs.

## Discussion

The current study examined whether cumulative or individual childhood experiences were more informative for understanding pain, subjective cognitive impairment, cognitive fatigue, and depression severity among adults with MS. Individual TRACEs and PCEs better explained each MS symptom than cumulative scores. Findings suggest that specific childhood experiences may provide meaningful information that is obscured when these experiences are aggregated into a cumulative score [24].

Findings provide support for the expansion of the traditional ACE framework to capture a broader range of childhood adversities, as postulated in the TRACE framework [24]. Nearly all participants (93.9%) reported at least one TRACE, and experiences outside the original ACE framework were among the most important predictors of MS symptoms. For example, racial/ethnic discrimination was one of the strongest predictors of pain and cognitive fatigue, natural disaster exposure was among the strongest predictors of pain and subjective cognitive impairment, and sudden loss of a loved one was the strongest individual predictor of both subjective cognitive impairment and cognitive fatigue. Thus, the TRACEs framework may offer a more comprehensive developmental framework for examining how childhood experiences relate to health in MS.

Findings also provide partial support for a life-course model in which childhood experiences contribute to later health through cumulative and potentially distinct developmental pathways. Greater cumulative TRACEs were associated with greater pain, subjective cognitive impairment, cognitive fatigue, and depression, consistent with the possibility that repeated exposure to adversity contributes to long-term vulnerability through interacting biological, psychological, and social processes [5]. However, the substantially stronger performance of individual-experience models indicates that accumulation alone does not fully characterize this process. Different childhood experiences were relatively more important for different MS symptoms, suggesting that the developmental consequences of adversity may depend on the type and context of exposure rather than solely on the number of adverse experiences. This distinction may be particularly important in MS, where symptom burden is heterogeneous and influenced by interacting neurological, psychological, and social factors.

Findings also highlight the importance of PCEs. Having at least one safe caregiver was among the strongest predictors across pain, subjective cognitive impairment, cognitive fatigue, and depression, and opportunities for enjoyment were among the more important predictors of depression. Greater PCEs were also associated with lower depression in correlational analyses. These findings are consistent with research demonstrating that PCEs are associated with better adult mental health and may promote resilience even among individuals exposed to adversity [14]. However, cumulative PCEs did not meaningfully improve prediction of pain, subjective cognitive impairment, or cognitive fatigue, and the TRACE and PCE interaction did not improve model performance. Thus, our findings do not support a simple model in which positive experiences uniformly buffer the effects of adversity. Instead, particular positive experiences, especially the presence of a safe caregiver, may independently be relevant to later health. Future research should examine whether specific PCEs have distinct associations with MS outcomes and whether their effects operate through biopsychosocial pathways.

The current study has several limitations. Childhood experiences were assessed retrospectively and may be affected by recall bias. The TRACEs and PCE measures capture whether experiences occurred but not their timing, chronicity, severity, frequency, or developmental context. The current study is cross-sectional, so temporal precedence among variables cannot be established. Future longitudinal research should examine whether specific childhood experiences are associated with MS onset, disease progression, or symptom trajectories and should determine whether timing or chronicity alters these associations. Additionally, participants were recruited online and resided within the United States, which is not representative of the broader MS population. Replication in clinically recruited and more diverse samples is warranted, particularly given the importance of racial/ethnic discrimination in the present models.

These findings have implications for trauma-informed MS care. Clinicians may benefit from considering childhood adversity when developing a comprehensive understanding of patients’ symptom burden, particularly when pain, cognitive difficulties, fatigue, and depression are prominent. Importantly, assessment should extend beyond the traditional ACE checklist to include experiences such as discrimination, bullying, community-level adversity, natural disasters, and significant loss. Such assessment should not be used to attribute current MS symptoms to childhood experiences or to replace evaluation of neurological and psychosocial contributors to symptom burden. Rather, understanding patients’ developmental histories may help clinicians recognize factors that could inform trauma-informed mental healthcare and provide individualized symptom-management strategies and relevant referrals and resources.

## Conclusion

Positive and traumatic and adverse childhood experiences were meaningfully associated with multiple MS symptoms, but their relative importance varied substantially across symptoms.

Individual PCEs and TRACEs consistently provided greater predictive information than cumulative scores, suggesting that reducing childhood experiences to a single cumulative score may obscure clinically relevant heterogeneity. The prominence of experiences such as racial/ethnic discrimination, natural disasters, and significant loss further supports an expanded conceptualization of childhood adversity that extends beyond the traditional ACE framework. Together, these findings highlight the value of examining childhood adversity and positive experiences as distinct and multidimensional components of the life-course context in which MS symptoms develop and progress.

## Data Availability

Due to the sensitive nature of the data in this study (i.e., childhood and present participant zip codes), data from the current study will only be shared upon request at the discretion of the corresponding author

